# Characteristics of those most vulnerable to employment changes during the COVID-19 pandemic: a nationally representative cross-sectional study in Wales

**DOI:** 10.1101/2020.11.03.20225144

**Authors:** Benjamin J Gray, Richard G Kyle, Jiao Song, Alisha R Davies

**Affiliations:** Research and Evaluation Division, Public Health Wales, Cardiff, UK

**Author notes:** Corresponding Author Dr Benjamin J Gray, Research and Evaluation Division, Knowledge Directorate, Public Health Wales, Number 2 Capital Quarter, Tyndall Street, Cardiff, CF10 4BZ.

## Abstract

**Background:** The public health response to the SARS-CoV-2 (COVID-19) pandemic has had a detrimental impact on employment and there are concerns the impact may be greatest amongst the most vulnerable. We examined the characteristics of those who experienced changes in employment status during the early months of the pandemic.

**Methods:** Data was collected from a cross-sectional, nationally representative household survey of the working age population (18-64 years) in Wales in May/June 2020 (N=1,379). We looked at changes in employment and being placed on furlough since February 2020 across demographics, contract type, job skill level, health status and household factors. Chi-squared or Fisher’s tests and multinomial logistic regression models examined associations between demographics, subgroups and employment outcomes.

**Results:** Of our respondents 91.0% remained in the same job in May/June 2020 as they were in February 2020, 5.7% were now in a new job, and 3.3% experienced unemployment. In addition, 24% of our respondents reported being placed on furlough. Non-permanent contract types, individuals who reported low mental wellbeing and household financial difficulties were all significant factors in experiencing unemployment. Being placed on ‘furlough’ was more likely in younger (18-29 years) and older (60-64 years) workers, those in lower skilled jobs and from households with less financial security.

**Conclusion:** A number of vulnerable population groups were observed to experience detrimental employment outcomes during the initial stage of the COVID-19 pandemic. Targeted support is needed to mitigate against both the direct impacts on employment, and indirect impacts on financial insecurity and health.

**What is already known on this subject?:**

- The response to the current global pandemic caused by SARS-CoV-2 (COVID-19) is already having a significant impact on people’s ability to work and employment status.
- Emerging UK employment data has raised concerns about the disproportionate impact on specific demographic groups.

**What this study adds?:**

- Groups that reported higher proportions of being placed on furlough included younger (18-29 years) and older (50-64 years) workers, people from more deprived areas, in lower skilled jobs, and those from households with less financial security.
- Job insecurity in the early months of the COVID-19 pandemic was experienced more by those self-employed or employed on atypical or fixed term contract arrangements compared to those holding permanent contracts.
- To ensure that health and wealth inequalities are not exacerbated by COVID-19 or the economic response to the pandemic, interventions should include the promotion of secure employment and target the groups identified as most susceptible to the emerging harms of the pandemic.

## Introduction

Employment is a wider determinant of health, and the links between good employment and better health outcomes are well established [1,2]. The response to the current global pandemic caused by SARS-CoV-2 (COVID-19) is already having a significant impact on people’s ability to work and employment status.

Global estimates suggest that up to 25 million jobs could be lost as a result of the COVID-19 pandemic [3]. Typically, mass unemployment events disproportionately impact the younger and older age groups [4–6], and those with lower skills or underlying health conditions are at more risk of exiting the labour market in the longer term. Compared to other Western countries, the United States (US) and the United Kingdom (UK) have experienced more severe immediate labour market impacts [7,8]. The unemployment rate in the US was estimated to be 20 per cent in April 2020 [7], and the unemployment rate in the UK reached a three year high of 4.5 per cent in August 2020 [9].

More specifically, in the UK, a greater fall in working hours was experienced by younger workers and those without guaranteed work [10], whilst declines in earnings have been hardest felt by the most deprived [10] and ethnic-minority communities [10,11]. The introduction of economic interventions such as the Coronavirus Job Retention Scheme (also known as ‘furlough’) will moderate the rise in redundancies initially, but a significant rise in unemployment is inevitable [12]. Predictions have suggested that job losses will be greatest within the retail and hospitality sectors [13,14] and women, young people, and the lowest paid are at particular risk of unemployment in this COVID-19 recession [14].

Identifying the groups most vulnerable to changes in employment during the COVID-19 pandemic is important, to better develop and target the health, re-employment and social support needed to prevent a longer term detrimental impact on societal health [4]. Emerging UK research has raised concerns about the disproportionate impact on specific demographic groups [10,11,15], whilst also commenting on regional disparities [15], suggesting a need for different approaches in the post-pandemic recovery. We investigated the impact of COVID-19 on employment in the initial phases of the pandemic as well as observed differences by underlying health and household financial security in Wales.

## Methods

### Data Source

The data included in this study was collected from the *COVID-19 Employment and Health in Wales Study*, a nationally representative cross-sectional online household survey undertaken between 25 May 2020 and 22 June 2020. The Health Research Authority approved the study (IRAS: 282223).

### Participants

Individuals were eligible to participate if they were resident in Wales, aged 18-64 years, and in employment in February 2020. Those in full time education or unemployed were not eligible to participate.

### Sample size calculation

In order to ensure the sample was representative of the Welsh population a stratified random probability sampling framework by age, gender, and deprivation quintile was used. A target sample size of 1,250 working-age adults was set to provide an adequate sample across socio-economic groups. To achieve a sample size of 1,250, a total of 20,000 households were invited to participate. These invitation figures were based on the proportion of eligible working age households in Wales and informed by the most recent mid-year population estimates and UK Labour Force Survey projections (figures for 2017 [16,17]). The 20,000 sample included a main sample of 15,000 and a boosted sample of 5,000 of those in the lower deprivation quintiles to ensure representation from the most deprived populations.

### Recruitment

Each selected household was sent a survey pack containing an invitation letter and participant information sheet. The invitation asked the eligible member of the household with the next birthday to participate in the survey. It included instructions on how to access the online questionnaire, by entering a unique reference number provided in the letter. The letter highlighted the value of responding to the survey, that participation was voluntary, responses would be confidential and provided an email address and Freephone telephone number to contact for further information, to request to complete the questionnaire by an alternative method (telephone or postal) or to inform the project team that they did not wish to participate. Any individuals who informed the project team that they did not meet the inclusion criteria or opted out were removed from the reminder mailing, which was posted 10 days after the initial invitation.

In total, 1,019 responses were received from the 15,000 base sample (6.8% response rate) and 273 responses received from the booster sample (5.5% response rate) resulting in 1,382 respondents (6.9% overall response rate). The majority of the responses were online questionnaires (99.1%), with an additional six paper and six telephone questionnaires. During data cleaning, individuals who had not completed the question on employment contract were excluded from the study, leaving a final sample of 1,379 for analysis.

### Questionnaire Measures

The employment details were collected at the date of questionnaire completion in May/June 2020, and were at this point also retrospectively asked about their employment situation in February 2020. Questions on employment including contract type, rights, and wages were based on the Employment Precariousness Scale (EPRES; [18]) and data on job role and associated skill level was determined using the current Standard Occupational Classification for the UK (SOC 2020; [19]). Questions were asked on any employment changes experienced between February 2020 and May/June 2020, the outcomes of interest were: (i) same job; (ii) new job, covering new job with same employer, new job with new employer and becoming self-employed; and (iii) unemployment. In addition, respondents were also asked if they had been placed on furlough since February 2020.

Explanatory variables included: socio-demographics (gender, age group, and deprivation quintile assigned based on postcode of residence using the Welsh Index of Multiple Deprivation (WIMD [20])); individual self-reported health status included general health and pre-existing health conditions (defined using validated questions from the National Survey for Wales [21]) and mental wellbeing (determined using the Short Version of the Warwick Edinburgh Mental Wellbeing Score [22]). We determined low mental wellbeing as 1 standard deviation below the mean score. Household factors were also collected including income covering basic needs [18], and child/ren in household. More detailed information on the questionnaire variables is provided in Table 1.

**Table 1.** Measures for variables included in the national survey.

| Measure | Source | Classification |
| --- | --- | --- |
| **Employment Contract** | Employment Precariousness Scale (EPRES) [18] | **Permanent** (*Permanent contract*)<br>**Fixed Term** (*Fixed-term, 1 year or more; fixed term, between 6 and 12 months; fixed-term, less than 6 months*)<br>**Atypical** (*Temporary, no fixed term; zero-hours contract; do not have a contract*)<br>**Self-Employed** (*Self-employed*) |
| **Employment skill level (Hierarchy)** | Office for National Statistics – SOC 2020 [19] | **Employment Skill Level 4** (*Corporate managers and directors; Science, research, engineering and technology professionals; Health professionals; Teaching and other educational professionals; Business, media and public service professionals*)<br>**Employment Skill Level 3** (*Other managers and proprietors; Science, engineering and technology associate professionals; Health and social care associate professionals; Protective service occupations; Culture, media and sports occupations; Business and public service associate professionals; Skilled agricultural and related trades; Skilled metal, electrical and electronic trades; Skilled construction and building trades; Textiles, printing and other skilled trades*)<br>**Employment Skill Level 2** (*Administrative occupations; Secretarial and related occupations; Caring personal service occupations; Leisure, travel and related personal occupations; Community and civil enforcement occupations; Sales occupations; Customer service occupations; Process, plant and machine operatives; Transport and mobile machine drivers and operatives*)<br>**Employment Skill Level 1** (*Elementary trades and related occupations; Elementary administration and service occupations*) |
| **Pre-existing (health) condition** | National Survey for Wales [21] | Do you have any physical or mental health conditions or illnesses lasting or expected to last for 12 months or more?<br>**Yes/No/Not Sure** |
| **General health** | National Survey for Wales [21] | How is your health in general? Is it...<br>**Good or better general health** (*good, very good*)<br>**Not good general health** (*fair, poor, very poor*) |
| **Mental Wellbeing** | The Warwick-Edinburgh Mental Wellbeing Scales (short version) [22] | Raw scores were converted into metric scores and categorised as **Average** or **Low Mental Wellbeing**. |
| **Household Income** | Employment Precariousness Scale (EPRES) [18] | Does your total household income allow you to cover your basic needs? (food, shelter and warmth)<br>**Always covers basic needs** (*always*)<br>**Does not cover basic needs** (*most of the time, sometimes, rarely, never*) |
| **Child(ren) in household** | Internal Question | How many children live with you in the following age bands? (enter a number)<br>a) 0-1 year old; b) 2-4 years old; c) Primary school age (5-10 years old); d) Secondary school age (11-17 years old)<br>**No child(ren) in household** (*total is zero*)<br>**Child(ren) in household** (*total is one or more*) |

### Statistical analysis

Data analysis on changes in employment were performed on the full sample (N=1,379). Not all respondents answered the question on furlough and any individuals who answered ‘don’t know’ were also excluded from the furlough analysis, leaving a sub-sample of 1,159. To examine differences in employment outcomes across population groups we tested the relationships between changes in employment or furlough and the explanatory variables using Chi-squared tests or Fisher’s exact test respectively. Multinomial logistic regression models were used to identify characteristics associated with changes in employment. Binary logistic regression was performed to identify characteristics associated with furlough. These results are reported as adjusted Odds Ratios (aOR) and 95% confidence intervals. A p-value of <0.05 was considered statistically significant. To supplement our multinomial logistic regression analysis, we explored the relationship between employment changes and contract type further through computing predicted probabilities whilst setting the remaining variables to their central measures.

## Results

### Sample Demographics

For reference, the demographic (gender, age, deprivation quintile) details of our ‘working age’ sample are compared to the latest Welsh population (mid-year 2018 population estimates [17]) in Table 2. Although broadly representative overall, compared to the Welsh population, females and the older age groups are over represented in our sample.

**Table 2.** Survey Population and Welsh Population Estimates (mid-year 2018) Comparisons.

|  | Survey Population |  | Welsh Population |  |
| --- | --- | --- | --- | --- |
|  | n | % | n | % |
| All | 1379 |  | 1,856,853 |  |
| Males | 542 | 39.3 | 924,020 | 49.8 |
| Females | 823 | 59.7 | 932, 833 | 50.2 |
| Not Provided | 14 | 1.0 |  |  |
| 18-29 Years | 157 | 11.4 | 485,909 | 26.2 |
| 30-39 Years | 271 | 19.7 | 371,851 | 20.0 |
| 40-49 Years | 338 | 24.5 | 375,526 | 20.2 |
| 50-59 Years | 416 | 30.2 | 433,915 | 23.4 |
| 60-64 Years | 177 | 12.8 | 189,652 | 10.2 |
| Not Provided | 20 | 1.4 |  |  |
| Quintile 1 (High Deprivation) | 258 | 18.7 | 371,014 | 20.0 |
| Quintile 2 | 326 | 23.6 | 370,637 | 20.0 |
| Quintile 3 | 228 | 16.5 | 384,927 | 20.7 |
| Quintile 4 | 254 | 18.4 | 370,242 | 19.9 |
| Quintile 5 (Low Deprivation} | 313 | 22.7 | 360,033 | 19.4 |

### Changes in employment status

Our findings suggest that 91.0% of the Welsh working age population were in the same job in May/June 2020 as they were in February 2020, 5.7% were now in a new job, and 3.3% have experienced unemployment (Table 3). There was no statistically significant difference observed in changes in employment by gender, age or deprivation quintile demographics (Table 3). Changes in employment were more apparent in those employed on non-permanent contracts (p<0.001; Table 3), where job losses were experienced more by those employed on an atypical contract (12.1%), fixed-term contract (7.7%), and also those who were self-employed (9.3%) compared to those employed on permanent arrangements (1.8%; Table 3). Unemployment was higher among those reporting financial difficulties in meeting basic needs (6.3%) compared to 2.2% of those with no financial struggles (p<0.001; Table 3) and also in those experiencing poorer mental health outcomes (low mental wellbeing: 11.5% compared to average mental wellbeing: 2.5%; p<0.001; Table 3).

**Table 3.**
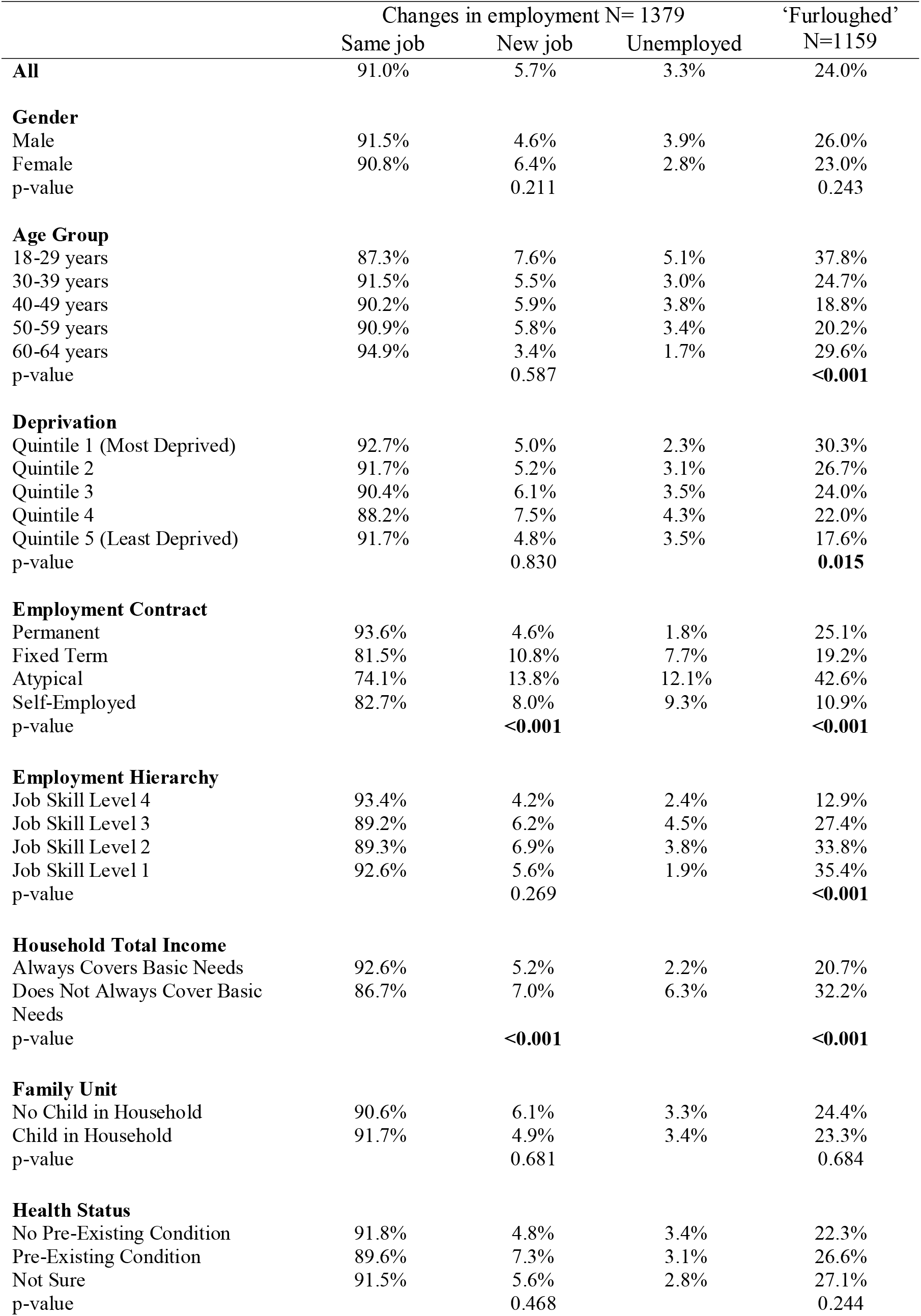
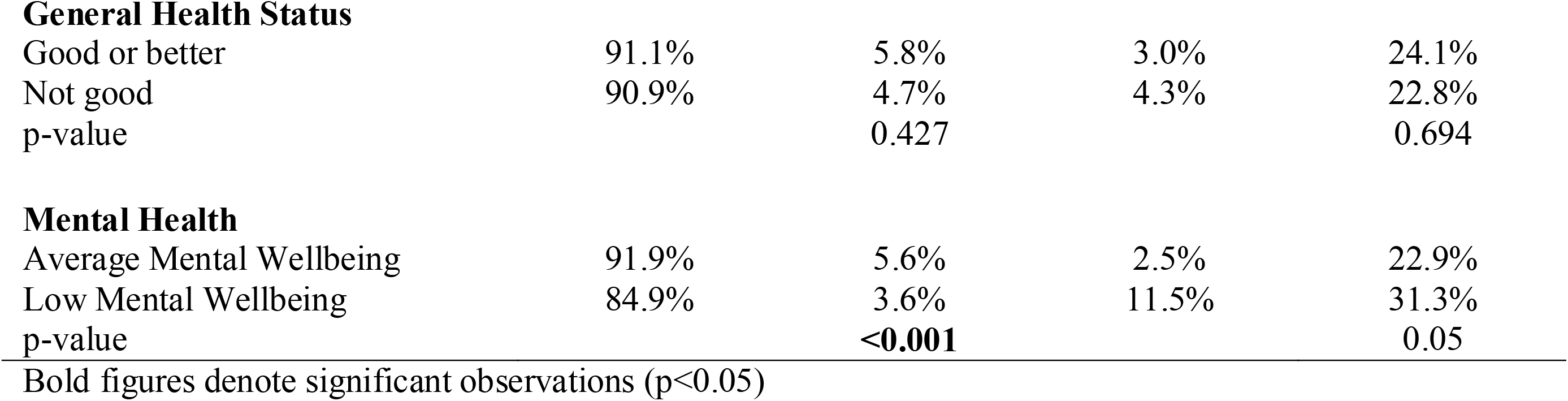
The Share of Employment Changes Experienced by Socio-Demographics, Wider Determinants, Health Status and Results of Chi-Squared Statistics.

|  | Changes in employment N= 1379 |  |  | 'Furloughed'<br>N=1159 |
| --- | --- | --- | --- | --- |
|  | Same job | New job | Unemployed |  |
| **All** | 91.0% | 5.7% | 3.3% | 24.0% |
| **Gender** |  |  |  |  |
| Male | 91.5% | 4.6% | 3.9% | 26.0% |
| Female | 90.8% | 6.4% | 2.8% | 23.0% |
| p-value |  | 0.211 |  | 0.243 |
| **Age Group** |  |  |  |  |
| 18-29 years | 87.3% | 7.6% | 5.1% | 37.8% |
| 30-39 years | 91.5% | 5.5% | 3.0% | 24.7% |
| 40-49 years | 90.2% | 5.9% | 3.8% | 18.8% |
| 50-59 years | 90.9% | 5.8% | 3.4% | 20.2% |
| 60-64 years | 94.9% | 3.4% | 1.7% | 29.6% |
| p-value |  | 0.587 |  | **<0.001** |
| **Deprivation** |  |  |  |  |
| Quintile 1 (Most Deprived) | 92.7% | 5.0% | 2.3% | 30.3% |
| Quintile 2 | 91.7% | 5.2% | 3.1% | 26.7% |
| Quintile 3 | 90.4% | 6.1% | 3.5% | 24.0% |
| Quintile 4 | 88.2% | 7.5% | 4.3% | 22.0% |
| Quintile 5 (Least Deprived) | 91.7% | 4.8% | 3.5% | 17.6% |
| p-value |  | 0.830 |  | **0.015** |
| **Employment Contract** |  |  |  |  |
| Permanent | 93.6% | 4.6% | 1.8% | 25.1% |
| Fixed Term | 81.5% | 10.8% | 7.7% | 19.2% |
| Atypical | 74.1% | 13.8% | 12.1% | 42.6% |
| Self-Employed | 82.7% | 8.0% | 9.3% | 10.9% |
| p-value |  | **<0.001** |  | **<0.001** |
| **Employment Hierarchy** |  |  |  |  |
| Job Skill Level 4 | 93.4% | 4.2% | 2.4% | 12.9% |
| Job Skill Level 3 | 89.2% | 6.2% | 4.5% | 27.4% |
| Job Skill Level 2 | 89.3% | 6.9% | 3.8% | 33.8% |
| Job Skill Level 1 | 92.6% | 5.6% | 1.9% | 35.4% |
| p-value |  | 0.269 |  | **<0.001** |
| **Household Total Income** |  |  |  |  |
| Always Covers Basic Needs | 92.6% | 5.2% | 2.2% | 20.7% |
| Does Not Always Cover Basic Needs | 86.7% | 7.0% | 6.3% | 32.2% |
| p-value |  | **<0.001** |  | **<0.001** |
| **Family Unit** |  |  |  |  |
| No Child in Household | 90.6% | 6.1% | 3.3% | 24.4% |
| Child in Household | 91.7% | 4.9% | 3.4% | 23.3% |
| p-value |  | 0.681 |  | 0.684 |
| **Health Status** |  |  |  |  |
| No Pre-Existing Condition | 91.8% | 4.8% | 3.4% | 22.3% |
| Pre-Existing Condition | 89.6% | 7.3% | 3.1% | 26.6% |
| Not Sure | 91.5% | 5.6% | 2.8% | 27.1% |
| p-value |  | 0.468 |  | 0.244 |
| **General Health Status** |  |  |  |  |
| Good or better | 91.1% | 5.8% | 3.0% | 24.1% |
| Not good | 90.9% | 4.7% | 4.3% | 22.8% |
| p-value |  | 0.427 |  | 0.694 |
| **Mental Health** |  |  |  |  |
| Average Mental Wellbeing | 91.9% | 5.6% | 2.5% | 22.9% |
| Low Mental Wellbeing | 84.9% | 3.6% | 11.5% | 31.3% |
| p-value |  | **<0.001** |  | 0.05 |

Bold figures denote significant observations (p<0.05)

### Characteristics of those furloughed

Considering demographics, the proportion of respondents placed on furlough was highest in the youngest age group (18-29 years; 37.8%), decreasing to 18.8% in the 40-49 years age group and increasing to 29.6% in the 60-64 years age group (p<0.001; Table 3). The highest proportion on furlough was evident amongst the most deprived communities (30.3%) and declined as a gradient across deprivation quintiles to 17.6% in the least deprived (p=0.015; Table 3).

Employment characteristics also impacted on being placed on furlough, lowest skill workers (35.4%) had the highest proportions ‘furloughed’ and this also decreased as a gradient with increasing skill level to 12.9% amongst the highest skilled workers (p<0.001; Table 3). People with atypical working arrangements experienced the highest proportions of being placed on furlough (42.6%; Table 3). A higher proportion of households struggling to cover basic financial needs also had been placed on furlough compared to those households reporting no financial difficulties (32.3% compared to 20.7%; p<0.001).

### Predictors of changes to employment situation and ‘furlough’

Younger people aged 18-29 years (aOR 2.5 [95% CI 1.5-4.3]) and older people aged 60-64 years (aOR 2.2 [95% CI 1.3-3.8]) were more likely to experience furlough compared to the 40-49 years age group (Table 4). Skill level was also a significant predictor of furlough, with those working in lower skilled roles more likely to have been placed on furlough compared to the highest skilled jobs (Job Skill 1: aOR 3.3 [95% CI 1.6-6.9]; Job Skill 2: aOR 3.2 [95% CI 2.2-4.7]; Job Skill 3: aOR 2.7 [95% CI 1.8-4.1]; Table 4). Individuals who experienced financial difficulties (aOR 1.9 [95% CI 1.4-2.6]) were also more likely to have been placed on furlough (Table 4). Those who were self-employed (aOR 0.3 [95% CI 0.2-0.6]) or who reported having ‘not good’ general health (aOR 0.6 [95% CI 0.4-0.9]) were less likely to have been placed on furlough (Table 4).

**Table 4.** Predictors of employment changes experienced in the early months of the COVID-19 pandemic. Data reported as adjusted Odds Ratios (aOR and 95% Confidence Intervals. Bold figures denote significant observations (p<0.05).

| Predictors | Change in employment status N=1379 |  | ‘Furloughed’<br>N=1159 |
| --- | --- | --- | --- |
|  | Now unemployed vs<br>Same job | New job vs Same job |  |
| **Gender** |  |  |  |
| Male | Reference | Reference | Reference |
| Female | 1.0 [0.5-2.0] | 1.4 [0.8-2.4] | 0.8 [0.6-1.2] |
| **Age Group** |  |  |  |
| 18-29 years | 1.3 [0.5-3.9] | 1.1 [0.5-2.6] | **2.5 [1.5-4.3]** |
| 30-39 years | 1.1 [0.4-2.9] | 1.0 [0.5-2.1] | 1.4 [0.9-2.3] |
| 40-49 years | Reference | Reference | Reference |
| 50-59 years | 0.7 [0.3-1.8] | 0.8 [0.4-1.5] | 1.3 [0.8-2.0] |
| 60-64 years | 0.4 [0.1-1.7] | **0.3 [0.1-0.9]** | **2.2 [1.3-3.8]** |
| **Deprivation** |  |  |  |
| Quintile 1<br>(Most Deprived) | 0.9 [0.3-2.7] | 0.9 [0.4-2.1] | 1.3 [0.8-2.1] |
| Quintile 2 | 1.4 [0.5-4.0] | 1.0 [0.5-2.2] | 1.3 [0.9-2.1] |
| Quintile 3 | 1.6 [0.6-4.7] | 1.3 [0.6-3.0] | 1.1 [0.7-1.9] |
| Quintile 4 | 1.8 [0.7-.5.1] | 1.9 [0.9-4.0] | 1.1 [0.7-1.8] |
| Quintile 5<br>(Least Deprived) | Reference | Reference | Reference |
| **Employment Contract** |  |  |  |
| Permanent | Reference | Reference | Reference |
| Fixed Term | **4.4 [1.3-14.8]** | **2.6 [1.1-6.3]** | 0.6 [0.3-1.3] |
| Atypical | **11.9 [4.3-32.9]** | **3.7 [1.5-9.1]** | 1.8 [0.96-3.3] |
| Self-Employed | **6.2 [2.7-14.1]** | 1.9 [0.9-4.1] | **0.3 [0.2-0.6]** |
| **Employment Hierarchy** |  |  |  |
| Job Skill Level 4 | Reference | Reference | Reference |
| Job Skill Level 3 | 1.6 [0.7-3.7] | 1.6 [0.8-3.0] | **2.7 [1.8-4.1]** |
| Job Skill Level 2 | 1.7 [0.7-3.9] | 1.8 [1.0-3.4] | **3.2 [2.2-4.7]** |
| Job Skill Level 1 | 0.4 [0.1-3.8] | 1.4 [0.4-5.0] | **3.3 [1.6-6.9]** |
| **Household Total Income** |  |  |  |
| Always Covers Basic<br>Needs | Reference | Reference | Reference |
| Does Not Always Cover<br>Basic Needs | **2.1 [1.1-4.3]** | 1.5 [0.9-2.6] | **1.9 [1.4-2.6]** |
| **Family Unit** |  |  |  |
| No Child in Household | Reference | Reference | Reference |
| Child in Household | 1.1 [0.5-2.4] | 0.7 [0.4-1.3] | 1.3 [0.9-1.8] |
| **Health Status** |  |  |  |
| No Pre-Existing Condition | Reference | Reference | Reference |
| Pre-Existing Condition | 0.7 [0.3-1.5] | **1.7 [1.0-3.07]** | 1.4 [1.0-1.9] |
| Not Sure | 0.5 [0.1-2.3] | 1.0 [0.3-3.4] | 1.1 [0.5-2.1] |
| **General Health Status** |  |  |  |
| Good or better | Reference | Reference | Reference |
| Not good | 1.2 [0.5-2.7] | 0.7 [0.4-1.5] | **0.6 [0.4-0.9]** |
| **Mental Health** |  |  |  |
| Average Mental Wellbeing | Reference | Reference | Reference |
| Low Mental Wellbeing | **4.1 [1.9-9.0]** | 0.6 [0.2-1.5] | 1.3 [0.8-2.2] |

Compared to permanent employment, the adjusted odds ratios (aOR) were distinctly higher for experiencing unemployment in all other contract types (atypical employment: aOR 11.9 [95% CI 4.3-32.9]; fixed-term contracts: aOR 4.4 [95% CI 1.3-14.8]; self-employed: aOR 6.2 [95% CI 2.7-14.1]; Table 4). In addition, those on atypical working arrangements (aOR 3.7 [95% CI 1.5-9.1]) and holding fixed-term contracts (aOR 2.6 [95% CI 1.1-6.3]) were more likely to have changed jobs. The computed predicted probabilities of falling into each of the three employment change categories were calculated among the different contract types (Table 5). These figures demonstrate further that job insecurity (changing jobs or becoming unemployment) is higher amongst those individuals holding non-permanent contracts. Furthermore, individuals who reported low mental wellbeing (aOR 4.14 [95% CI 1.9-9.0]) or experienced financial difficulties (aOR 2.14 [95% CI 1.1-4.3]) were also more likely to experience unemployment (Table 4).

**Table 5.** Predicted probabilities derived from multinomial logistic regression for employment changes experienced by contract type.

|  | Changes in employment N= 1379 |  |  |
| --- | --- | --- | --- |
|  | Same job | New job | Unemployed |
| **Employment Contract** |  |  |  |
| Permanent | 96.8% | 2.6% | 0.6% |
| Fixed Term | 90.9% | 6.3% | 2.8% |
| Atypical | 84.7% | 8.4% | 6.9% |
| Self-Employed | 91.5% | 4.6% | 3.9% |

## Discussion

This study reports findings from the first nationally representative survey in Wales that examines the associations between socio-demographics, wider determinants, underlying health status and employment outcomes during the COVID-19 pandemic. The findings provide unique insights into the population groups experiencing societal harms [23] as a result of the indirect effect of COVID-19 on employment. People who are younger (18-29 years), older (60-64 years), living in the most deprived communities, employed on non-permanent contracts, low skilled workers, and those with less financial security are more likely to experience employment harms as a result of the COVID-19 pandemic. Our study therefore identifies vulnerable groups that are ‘at risk’ of future jobs losses, and also reveals the disproportionate experiences of population subgroups in relation to unemployment experienced in the early part of the pandemic.

These findings are consistent with early evidence from other parts of the UK in relation to the at-risk populations that have been furloughed, notably, those in certain age groups (18-29 years and 60 years and older) and those in lower skilled jobs [13,14]. Of concern, however, is the disproportionate impact on vulnerable groups in the population that are currently supported by the Coronavirus Job Retention Scheme (‘furlough’). Not all individuals placed on furlough (and subsequent job retention schemes) will ultimately lose their jobs, but there is the potential for the impact on employment and health to be greatest amongst the most vulnerable sub-populations when this scheme ceases [12]. Evidence indicates that pandemics have the potential to exacerbate inequalities [6,24], especially within the most deprived communities and our findings suggest COVID-19 will have a similar impact. One of the more striking observations is the unequal impacts of employment changes on those people employed on non-permanent contract arrangements. Existing research from the early months of the pandemic has also reported that those with temporary contracts were more likely to have experienced unemployment as a result of the coronavirus shock [8]. In recent decades, employment trends have seen a marked increase in flexible, non-standard arrangements; contributing to reduced job-security reduced income-security, and increased temporary contracts [25,26]. It is well documented that these precarious employment arrangements are more commonplace within younger, migrant and female subpopulations, and there is growing evidence to suggest there are negative impacts on health [26,27]. Those on atypical and fixed-term contracts were also more likely to have changed jobs since February 2020, longitudinal research is required to assess the quality of this new employment and the potential longer-term implications on health.

Unemployment is also known to have a negative impact on an individual’s own health, such as poorer mental health outcomes [28,29]. Our data confirms this association. This worrying finding warrants further investigation and intervention as, although causality cannot be established through our study, it may reflect a consequence of unemployment or furlough during the pandemic rather than a pre-existing state. However, research has suggested that mental health in the UK has deteriorated compared to pre-COVID trends [30]. Being, or in the case of our study, becoming unemployed during a recession can worsen levels of psychological distress [31,32]. Our findings also suggest that those with pre-existing health conditions disproportionately experienced job loss in the early part of the pandemic. This echoes a pre-COVID European study where those with poorer mental and physical health were at greater risk of job losses [33]. Addressing poorer health outcomes associated with poverty was already a public health priority before the COVID-19 pandemic [34,35]. Our results suggest households struggling financially to meet basic needs have been disproportionately impacted by unemployment during the early part of the pandemic, and this may have potential to cause wider harm to other members in the household [36,37].

Our study helps to inform strategies and interventions to support vulnerable groups who have already disproportionately experienced harm from the early part of the pandemic and more importantly, re-emphasises the importance of permanent contract arrangements to negate adverse impacts of economic shocks. Uncertainties surrounding the global post-COVID labour market remain and although job retention schemes in place in many countries across the world still have some months to run these are economic rather than health-driven solutions. The potential for long-term negative impacts on health and wellbeing is evident in our study and health-aligned solutions may be required to mitigate these negative consequences. It is also important to remember that job insecurity itself, even if only perceived, can also have negative health consequences [38,39]. Furthermore, given poverty and health are inextricably linked [34–37], the higher levels of furlough we observed among households who reported struggling financially to cover basic needs, requires attention. Social support systems and targeted initiatives to address inequalities in access to the labour market are needed by those potentially facing unemployment. Our study underscores the need to draw public health professionals and practices into the heart of debates around economic recovery and restructuring to ensure wider determinants of health and health inequalities are addressed [40].

### Study limitations

Our study has three main limitations. First, the cross-sectional design of the survey means that the observations demonstrate an association rather than causality. For example, caution is needed in interpretation of some of the findings in relation to mental wellbeing due to the data collection being at one time point and it is not known if low mental wellbeing was evident before. As noted, it has been observed that trends in UK mental health have worsened from pre-COVID levels [30]. Second, employment changes were a relatively rare event during the early stages of the pandemic, although this manuscript clearly demonstrates some important findings, some of the adjusted odds ratios should be interpreted with caution. To this end, for a more nuanced interpretation we included predicted probabilities of falling into each of the three employment change status among people holding different types of contracts. Despite the low likelihood of job loss, employees on atypical contrasts are at increased risk over other types of contracts. Finally, although designed to be representative to the population, females and the older age groups are over represented in our sample compared to the Welsh population, whereas deprivation quintiles are broadly representative except for the middle-to-high quintiles (Quintiles 3 and 4). However, the consistencies within our data and national data (where comparators are available) suggest that our findings are generalisable. Future studies that examine the longer-term impacts of COVID-19 on employment and health could adopt a household door-to-door approach (if restrictions allow) to improve response rate and representativity.

## Conclusion

Unemployment in the early months of the COVID-19 pandemic impacted most on individuals in non-permanent work and those experiencing poorer mental wellbeing or financial difficulties. Furlough disproportionately impacted several population groups including the youngest (18-29 years) and oldest (60-64 years) age groups, people living in deprived communities, those employed in lower skilled job roles, and people struggling financially. A social gradient was observed across deprivation and worker skill level with those living in the most deprived areas and working in the lowest skilled jobs more likely to be furloughed. Interventions to support economic recovery need to target the groups identified here as most susceptible to the emerging harms of the pandemic. Our study also strongly emphasises the importance of good, secure employment to survive economic shocks and protect individuals from the negative harms of unemployment.

## Data Availability

The data presented in this manuscript is part of the 'COVID-19, Employment and Health in Wales' study and available only to the research team.

## Conflicts of Interest

The authors wish to declare no competing interests.

## Acknowledgements

The authors express their gratitude to MEL Research who completed the data collection for this study and to the people from across Wales who completed the survey. We would also like to acknowledge the contribution of our colleague James Bailey for his assistance in the initial stages of the manuscript.

## Notes

### Competing Interest Statement

The authors have declared no competing interest.

### Funding Statement

This study received no external funding.

### Author Declarations

The Health Research Authority approved the study (IRAS: 282223).

### Summary of Updates

Revised manuscript uploaded to reflect reviewer comments.

